# Efficacy and Safety of Hydroxychloroquine and Chloroquine for COVID-19: A Systematic Review

**DOI:** 10.1101/2020.05.19.20106906

**Authors:** Sonal Singh, Thomas J. Moore

## Abstract

**BACKGROUND:** Hydroxychloroquine and chloroquine are widely used to treat hospitalized COVID-19 patients primarily based on antiviral activity in *in vitro* studies. Our objective was to systematically evaluate their efficacy and safety in hospitalized patients with COVID-19.

**METHODS:** We systematically reviewed PubMed, ClinicalTrials.gov, and Medrxviv for studies of hydroxychloroquine and chloroquine in COVID-19 hospitalized patients on April 26, 2020. We evaluated the quality of trials and observational studies using the Jadad criteria and Newcastle Ottawa Scale, respectively.

**RESULTS:** After a review of 175 citations, we included 5 clinical trials (total of 345 patients), 9 observational studies (n = 2529), and 6 additional studies (n = 775) reporting on the QT interval. Three studies reported treatment benefits including two studies reporting benefit on virologic outcomes, which was statistically significant in one study, and another reported significant improvement on cough symptoms. Three studies reported that treatment was potentially harmful, including an significantly increased risk of mortality in two studies and increased need for respiratory support in another. Eight studies were unable to detect improvements on virologic outcomes (n = 3) or pneumonia or transfer to ICU/death (n = 5). The proportion of participants with critical QTc intervals of ≥ 500 ms or an increase of ≥ 60 ms from baseline ranged from 8.3% to 36% (n = 8). One clinical trial and six observational studies were of good quality. The remaining studies were of poor quality.

**CONCLUSIONS:** Our systematic review of reported clinical studies did not identify substantial evidence to support the efficacy of hydroxychloroquine or chloroquine in hospitalized COVID-19 patients and raises questions about potential harm from QT prolongation and increased mortality.

Key Points
- We conducted a systematic review of the efficacy and safety of hydroxychloroquine and chloroquine among patients hospitalized with COVID-19 and identified 14 studies reporting on clinical or virologic outcomes and 6 additional studies reporting on the QT interval.
- We conducted a systematic review of the efficacy and safety of hydroxychloroquine and chloroquine among patients hospitalized with COVID-19 and identified 14 studies reporting on clinical or virologic outcomes and 6 additional studies reporting on the QT interval.
- Hydroxychloroquine or chloroquine improved virologic outcomes in 2 clinical studies and cough in another study.
- We conducted a systematic review of the efficacy and safety of hydroxychloroquine and chloroquine among patients hospitalized with COVID-19 and identified 14 studies reporting on clinical or virologic outcomes and 6 additional studies reporting on the QT interval.
- Hydroxychloroquine or chloroquine improved virologic outcomes in 2 clinical studies and cough in another study.

## INTRODUCTION

There are no known effective drugs for acute respiratory syndrome coronavirus-2 (SARS-CoV-2). Hydroxychloroquine and chloroquine raise the endosomal pH required for virus/cell fusion and interfere with the glycosylation of SARS-corona virus to the cell-surface receptor.^1^ Chloroquine is a potent inhibitor of SARS-CoV-1 coronavirus infection in primate cell cultures,^2^ and both have antiviral activity *in vitro* against SARS-CoV-2.^3 4^ They also have adverse effects, including prolongation of the QT interval,^5^ retinal damage, psoriasis outbreaks, myopathy, and suicidal behaviors.^6^ The two drugs are inexpensive and approved for prevention of malaria and treatment of lupus and rheumatoid arthritis.

The two drugs were heralded as a “breakthrough” treatment based on preliminary data,^7^ and their use grew exponentially in patients with COVID-19. A global survey of 6150 physicians reported that 55% had used hydroxychloroquine.^8^ Monthly average outpatient hydroxychloroquine prescriptions in the United States increased from 15,000 to 139,000 in March 2020 compared to prior years.^9^ The U.S. Food and Drug Administration (FDA) granted an emergency use authorization for treatment of the virus in hospitalized patients,^10^ and made supplies from the national emergency stockpile available to hospitals despite noting that dose and duration were unknown. Although several international agencies recommended use,^7^ an National Institutes of Health panel did not find sufficient evidence to recommend for or against use.^11^ In April 2020, the FDA issued a Drug Safety Communication warning of the cardiovascular risks of the two drugs.^12^ Our objective was to systematically evaluate the efficacy of hydroxychloroquine and chloroquine in hospitalized patients with COVID-19.

## METHODS

### Data Sources and Systematic Search Strategy

We searched PubMed, ClinicalTrials.gov, and Medrxviv for studies of patients hospitalized with COVID-19 and treated with hydroxychloroquine, chloroquine, and/or combinations with antibiotics. We initially searched PubMed for studies published from November 1, 2019, to April 26, 2020. From Clinicaltrials.gov, we extracted studies with results reported as of April 26, 2020. We also searched for preprints at medrxviv.com with similar terms and signed up to receive electronic notification of new articles through May 10, 2020. We reviewed the references of included studies for additional studies. Our search strategy is shown in the **Supplementary Table S1**.

### Eligibility Criteria

We included both published and unpublished clinical trials and observational studies that reported on chloroquine and hydroxychloroquine use either as a single drug or in combination with azithromycin to treat patients hospitalized patients with COVID-19. We excluded case reports and *in vitro* studies.

### Outcomes

The included clinical studies reported on any of the following outcomes: virologic clearance as measured by polymerase chain reaction (PCR) or culture, symptom improvement *(e.g*., cough), improvement in radiological assessment of pneumonia, hospital discharge and need for intubation or respiratory support or death. Additionally, we analyzed separately studies reporting on QT prolongation.

### Study Selection

Two reviewers evaluated the citations to identify relevant studies. We resolved discrepancies through discussion after review of full texts and achieved full agreement prior to inclusion.

### Data Extraction

We extracted data on study design, location, number of hospitals, outcomes, interventions, and control including dose and duration of the study drug and measures of statistical significance.

### Quality Assessment

We evaluated the quality of clinical trials using the Jadad criteria^13^ and the quality of observational studies using the Newcastle Ottawa Scale.^14^ Controlled observational studies were classified as being good, fair, or poor based on their performance on measures of selection, comparability, and outcomes. We rated all uncontrolled studies as being of poor quality.

### Data Synthesis

We conducted a qualitative synthesis of the evidence since the data were too heterogeneous to be pooled in a meta-analysis.

## RESULTS

### Search Results

The results of our search are shown in the flow sheet in **Figure 1**.

**Figure 1.**
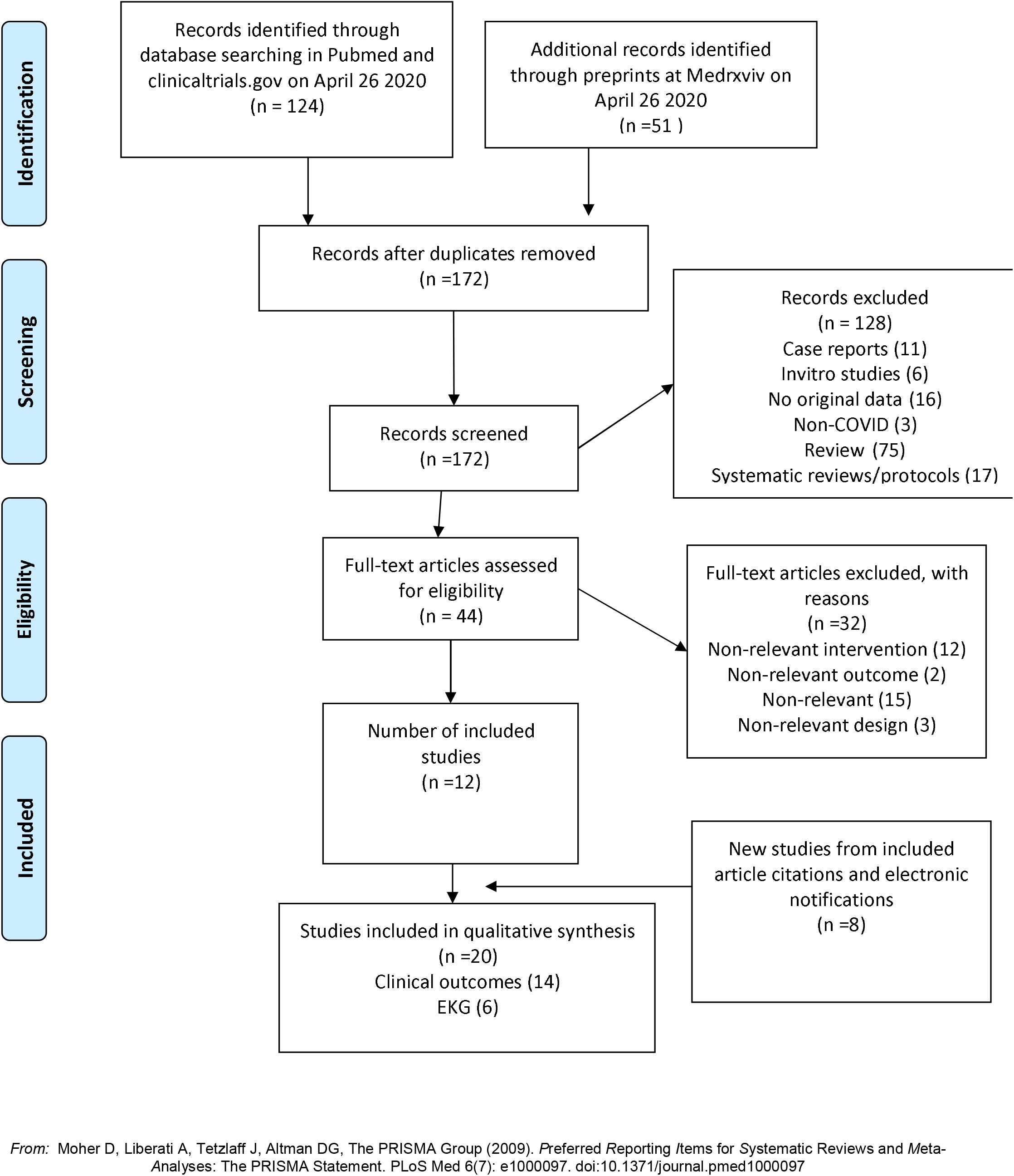
Flow Sheet of Included Studies

Our search identified 175 citations. After a review of citations, we included 5 clinical trials^15-19^ 7 controlled observational studies,^20-26^ and 2 uncontrolled observational studies on clinical outcomes.^27 28^ Six uncontrolled studies reporting on changes in the QT interval were also evaluated separately.^29-34^ In case of duplicate publication, we extracted data from the most recent study.^29 35^

### Study Characteristics

The characteristics of clinical trials and observational studies are shown in **Table 1**. Four clinical trials were conducted in China^15-17 19^ and one in Brazil.^18^ The total number of participants enrolled in the trials was 345 and the sample size of clinical trials ranged from 22^19^ to 150.^15^ Three clinical trials evaluated hydroxychloroquine vs usual care,^15-17^ one evaluated chloroquine vs lopinavir/ritonavir, and another evaluated high-dose chloroquine (600 mg twice daily for 10 days) vs low-dose chloroquine (450 mg twice daily for 5 days).^18^

**Table 1.** Clinical studies of chloroquine and hydroxychloroquine in patients hospitalized with COVID-19

| Study | Country | Patients, number | Hospitals, number | Treatment | Comparator |
| --- | --- | --- | --- | --- | --- |
| **Randomized controlled Trials** |  |  |  |  |  |
| Borba et al 2020 <sup>18</sup> | Brazil | 81 | 1 | CQ | Low dose CQ |
| Chen et al (a) 2020 <sup>16</sup> | China | 62 | 1 | HCQ | Usual care |
| Chen et al (b) 2020 <sup>17</sup> | China | 30 | 1 | HCQ | Usual care |
| Huang et al 2020 <sup>19</sup> | China | 22 | 1 | HCQ | Lopinavir/ritonavir |
| Tang et al 2020 <sup>15</sup> | China | 150 | 16 | HCQ | Usual care |
| **Observational studies** |  |  |  |  |  |
| Barbosa et al 2020 <sup>25</sup> | USA | 63 | 2 | HCQ | Usual care |
| Chen et al (c) 2020 <sup>24</sup> | China | 284 | 1 | CQ | Usual care |
| Feng et al 2020 <sup>23</sup> | China | 50 | 1 | CQ | Usual care |
| Gautret et al 2020 (a) <sup>22</sup> | France | 46 | 4 | HCQ | Usual care |
| Geleris et al 2020 <sup>26</sup> | US | 1446 | 1 | HCQ | Usual care/matched cohort |
| Mahevas et al 2020 <sup>20</sup> | France | 181 | 4 | HCQ | Usual care |
| Magagnoli et al 2020 <sup>21</sup> | US | 368 | 170 | HCQ/<br>HCQ+AZ | Males, matched cohort>65 yrs |
| Gautret et al (b) 2020 <sup>28</sup> † | France | 80 | 1 | HCQ+AZ | None |
| Molina et al 2020 <sup>27</sup> † | France | 11 | 1 | HCQ+AZ | None |
| † Uncontrolled observational studies. The remaining observational studies were controlled. |  |  |  |  |  |
| Abbreviations: AZ=Azithromycin; CQ=Chloroquine; HCQ=Hydroxychloroquine, |  |  |  |  |  |

Among the nine observational studies reporting on clinical outcomes, four were conducted in France,^20 22 27 28^ three in the United States,^25 21 26^ and two in China.^23 24^ The sample size of observational studies ranged from 11^27^ to 1446.^26^ Two studies reported on the use of chloroquine,^23 24^ while the remaining studies evaluated either hydroxychloroquine alone^21 22 25^ or in combination with azithromycin.

Among the six studies exclusively reporting on the QT interval as an outcome, three were conducted in the United States,^30 33 34^ one in France,^32^ and another in the Netherlands.^31^ In addition, one multicenter study was conducted in the United States and Italy.^29^ Five studies evaluated hydroxychloroquine + azithromycin,^29 30 32-34^ and one evaluated chloroquine alone.^31^ One study evaluated all three drugs.^34^

### Study Quality

The quality assessments of clinical trials and observational studies is shown in **Supplementary Figures S1 and S2**, respectively. Randomization was adequately reported in only two clinical trials,^15 18^ whereas the remaining trials did not provide adequate details of randomization.^16 17 19^ One trial was appropriately double masked,^18^ and another reported that neither the investigators or participants were aware of treatment assignment, but did not describe details of blinding. ^16^One trial was open label in which neither the participants, investigator or outcome assessors were blinded,^15^ and the remaining three trials did not report on blinding., ^17 19^ Two trials reported on dropouts.^17 18^ Only three clinical trials were peer reviewed.^17-19^ Two trials reported an intention to treat analysis.^17 18^

Among nine observational studies reporting on clinical outcomes, six controlled observational studies were rated as being of good quality,^20 21 23-26^ whereas one controlled observational study, ^22^ and the two uncontrolled studies,^27 28^ were of poor quality. Four studies were peer reviewed,^22 26-28^ five were not.^20 21 23-25^

All six uncontrolled studies reporting on QT interval alone were rated as being of poor quality,^29-34^ and four were peer reviewed.^31-34^

### Results on Benefits and Harms

The results on potential benefits and harms is shown in **Table 2**. Among those reporting potential benefits, two observational studies reported beneficial effects on virologic clearance with the combination of azithromycin and hydroxychloroquine,^22 28^ and one clinical trial reported a statistically significant benefit in a reduction in cough remission time.^17^ Three studies reported an increase in potential harms including increasing need for respiratory support^25^ and increasing mortality with high-dose chloroquine^18^ and hydroxychloroquine.^21^ The remaining studies did not detect any statistically significant difference in either virologic clearance,^16 19 24 27^, pneumonia,^17 19 23^ transfer to ICU or death,^20 26^ or intubation or death.^26^

**Table 2.** Virologic and clinical outcomes for hydroxychloroquine and chloroquine in patients hospitalized with COVID-19

| Study | Number on active drug | Active drug | Daily dose, mg | Treatment duration (days) | Endpoint | Study quality |
| --- | --- | --- | --- | --- | --- | --- |
| **POTENTIAL BENEFITS** |  |  |  |  |  |  |
| **Virologic outcomes** |  |  |  |  |  |  |
| Gautret et al (a) 2020 <sup>22</sup> | 26 | HCQ+AZ | 600 | 10 | ↑Virologic clearance<sup>†</sup> | Poor |
| Gautret et al (b) 2020 <sup>28</sup> | 80 | HCQ+AZ | 600 | 10 | ↑Virologic clearance | Poor |
| **Clinical outcomes** |  |  |  |  |  |  |
| Chen et al (b) 2020 <sup>17</sup> | 15 | HCQ | 400 | 5 | ↓Cough remission time<sup>†</sup> | Poor |
| **POTENTIAL HARMS** |  |  |  |  |  |  |
| **Respiratory support & mortality** |  |  |  |  |  |  |
| Barbosa et al 2020 <sup>25</sup> | 32 | HCQ | 800,200-400 | 1-2, 3-4 | ↑ Respiratory support<sup>†</sup> | Good |
| Borba et al 2020 <sup>18</sup> | 41 | CQ | 1200 | 10 | ↑ Mortality<sup>†</sup> | Good |
| Magagnoli et al 2020 <sup>21</sup> | 97,113 | HCQ, HCQ+AZ | NR | NR | ↑ Mortality<sup>†</sup> | Good |
| **NO BENEFITS DETECTED** |  |  |  |  |  |  |
| **Virologic outcomes** |  |  |  |  |  |  |
| Tang et al 2020 <sup>15</sup> | 75 | HCQ | 1200, 800 | 3,9-19 | Virologic clearance & symptoms | Poor |
| Chen et al (c) 2020 <sup>24</sup> | 25 | CQ | NR | NR | Virologic clearance | Good |
| Molina et al 2020 <sup>27</sup> | 11 | HCQ+AZ | 600 | 10 | Virologic clearance | Poor |
| **Clinical outcomes and death** |  |  |  |  |  |  |
| Chen et al (a) 2020 <sup>16</sup> | 31 | HCQ | 400 | 5 | Pneumonia | Poor |
| Huang et al 2020 <sup>19</sup> | 10 | CQ | 1000 | 10 | Pneumonia & virologic clearance | Poor |
| Feng et al 2020 <sup>23</sup> | 25 | CQ | 1000 | NR | Pneumonia | Good |
| Mahevas et al 2020 <sup>20</sup> | 84 | HCQ | 600 | 7 | Transfer to ICU/death | Good |
| Geleris et al 2020 <sup>26</sup> | 811 | HCQ | 1200, 400 | 5 | Intubation/death | Good |
| †Statistically significant compared to control. |  |  |  |  |  |  |
| Abbreviations: AZ=Azithromycin; CQ=Chloroquine; HCQ=Hydroxychloroquine, NR=Not Reported |  |  |  |  |  |  |

### Virologic Clearance

One controlled observational study reported statistically significantly increased virologic clearance with hydroxychloroquine compared to controls (n = 26, 70% vs 12%; p<0.001); and with hydroxychloroquine + azithromycin vs controls (n = 6, 100% vs 12%; p<0.001).^22^ An uncontrolled observational study from the same hospital which included a few participants from the earlier study reported that 93% of 80 initial participants were PCR negative at Day 8.^28^ In contrast, there was no statistically significant difference on virologic clearance in two clinical trials that evaluated hydroxychloroquine against controls,^15 16^ and in one clinical trial^10^ and another observational study^24^ that evaluated chloroquine and controls.^19 24^ Another uncontrolled study reported low rates of virologic clearance in 2/10 (20%) participants on hydroxychloroquine + azithromycin.^27^

### Clinical Outcomes

One trial enrolling 30 patients reported statistically significant improvement in remission of symptoms of cough with hydroxychloroquine compared to controls,^16^ although there was no difference in symptom improvement in one trial, ^15^ or clinical recovery in another.^19^ Borba *et al* reported no significant difference in the need for mechanical ventilation (20% vs 10.5%; p = 0.41) at Day 6 between high-dose and low-dose chloroquine arms in a clinical trial.^18^ Barbosa *et al* reported a statistically significant increased need for respiratory support with hydroxychloroquine compared to the usual care in both the overall cohort and the matched sub-cohort,^25^ Mahevas *et al* reported no significant difference in the need for transfer to ICU or death between hydroxychloroquine and usual care (RR 0.93; 0.48-1.81) in an observational study.^20^ Magagnoli *et al* reported no significant difference in the need for ventilation when comparing vs control either hydroxychloroquine (adjusted Hazard Ratio [aHR] 1.43; 95% CI 0.53-3.79) hydroxychloroquine + azithromycin (aHR 0.43; 95% CI 0.16-1.12) respectively in an observational study.^21^ In the largest propensity matched database study conducted in the US,^26^ patients on hydroxychloroquine were more likely to have the primary endpoint of intubation or death in the unadjusted analysis (HR 2.37; 95% CI 1.84-3.02), but there was no statistically significant difference between hydroxychloroquine and control in the main analysis using inverse probability weighting according to propensity score (aHR 1.04; 95% CI 0.82-1.32). Three studies reported no significant difference in the risk of pneumonia between chloroquine and controls.^16 19 23^ There was no statistically significant difference in either hospital stay in one observational study, ^24^ or in hospital discharge in a clinical trial.^19^

### Mortality

Borba *et al* compared high-dose chloroquine to low-dose chloroquine and the trial was prematurely halted because of statistically significantly increased mortality in the high-dose chloroquine arm (39% vs 15%; Odds Ratio 3.6, 95% CI, 1.2-10.6).^18^ Magagnoli *et al* also reported a statistically significantly increased risk of mortality with hydroxychloroquine compared to control (aHR 2.61; 95% CI 1.1-6.17), but not for hydroxychloroquine + azithromycin (aHR 1.14; 95% CI 0.56-2.32).^21^ Geleris *et al* reported an unadjusted excess of deaths among participants in the hydroxychloroquine arm compared to controls (157/811 vs 75/565; 19.3% vs 13.3%) without conducting any formal tests of statistical significance, although hydroxychloroquine treated patients were more severely ill at baseline.^26^ One small trial reported no deaths among any participants at the end of the study,^17^ whereas the remaining controlled studies reported small numbers of deaths without meaningful difference between groups.^20 22 25^

### QT Prolongation

A critical QTc interval ≥ 500 ms or an increase ≥ 60 ms was noted in a clinical trial,^18^ a controlled observational study,^20^ and six uncontrolled studies,^29-34^ with proportions of treated patients affected ranging from 8.3%^20^ to 36% as shown in **Figure 2**.^32^

**Figure 2.**
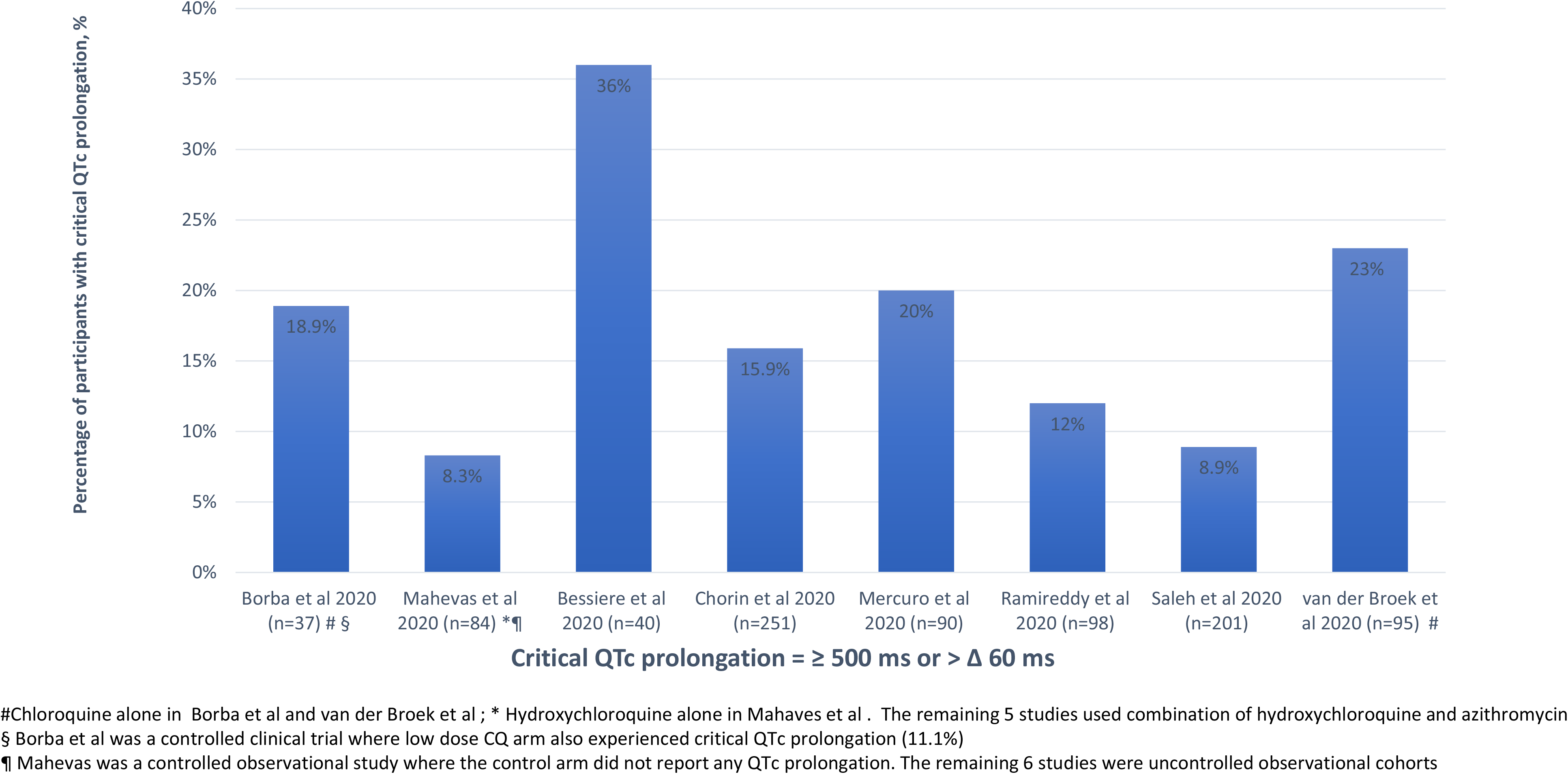
Critical QTc Prolongation in Studies of Chloroquine/ Hydroxychloroquine in Patients with COVID-19

Borba *et al* showed a statistically significant higher risk of QTc ≥ 500 ms in the high-dose chloroquine arm compared to low-dose chloroquine (18.9% vs 11.1%),^18^ and Mahevas *et al* reported that a higher number of participants on hydroxychloroquine experienced QTc prolongation ≥ 60 ms compared to none in controls (8.3% vs 0%).^20^

The combination of hydroxychloroquine + azithromycin caused a statistically significantly increased risk of QT prolongation compared with hydroxychloroquine alone in four studies.^31-34^ Bessiere *et al* (n = 40) reported 33% vs 5% (p = 0.03) experienced QTc ≥ 500 ms for the combination with azithromycin vs hydroxychloroquine alone and also reported that 93% of patients in both groups combined had QT prolongation compared to baseline.^32^ Mercuro *et al* (n = 90) reported a statistically significant median (Interquartile range [IQR]) increase in QT interval in the combination of hydroxychloroquine + azithromycin compared to hydroxychloroquine alone (23 seconds; (IQR) [10 seconds - 40 seconds] vs 5.5 seconds; IQR [-15.5 seconds to 34.25 seconds]; P=.03).^33^ Saleh *et al* reported that the maximum QTc was significantly longer in the combination vs monotherapy group (n = 201; 470.4 ms +/- 45 ms vs 453.3ms +/-37 ms; p = 0.004),^34^ although critical QTc > 500 ms was not different between the groups. Van den Broek *et al* (n=95) reported that the mean QTc prolongation was 34 ms (95% CI 25-43 ms).^31^

Two studies reported the occurrence of *torsades de pointes* on treatment,^29 33^ whereas six studies specifically reported the absence of *torsades*.^25 30-32 34 35^ Borba *et al* reported that two of the participants in the high-dose chloroquine experienced ventricular tachycardia compared to no participants in the low-dose arm.^18^

## DISCUSSION

Our systematic review shows that 11/14 (78.6%) of the clinical studies either reported evidence of increased harms (n = 3) or detected no beneficial effects (n = 8). In the studies reporting treatment benefits, efficacy was mainly limited to effects on virologic clearance (n = 2 studies) or cough (n=1 study), without detecting improvements in the risk of pneumonia, need for mechanical ventilation, or mortality. In contrast, the safety data raise concerns about an increased need for respiratory support,^25^ mortality,^18 21^ and critical QT prolongation.^18 20 29-34^

The beneficial effects on virologic clearance reported from two studies,^22 28^ need to be weighed against the lack of effect on virologic outcomes from several other studies,^15 17 19 24 27^ and those reporting potential harms.^18 20 21^ Also, one of the studies reporting increased virologic clearance^22^ was later deemed not to meet the International Society of Antimicrobial Chemotherapy expected journal standards.^36^ An independent Bayesian re-analysis concluded that the study was unable to determine the clinical effect of hydroxychloroquine on SARS-CoV-2 viral load reduction because it did not compute the effect of monotherapy against controls.^37^ We noted critical QTc prolongation in all eight studies reporting on this endpoint,^18 20 29-34^ with four studies reporting that the adverse effects increased with concomitant azithromycin therapy.^31-34^ This new group of studies advances the strength of the evidence implicating hydroxychloroquine since the prescribing information cites only an unspecified number of postmarketing reports.^6^

Our systematic review should be distinguished from the results of existing systematic reviews.^38-43^ One review included *in vitro* studies, editorials, opinion letters, expert consensus, and guidelines.^39^ Shah *et* al^41^ included only one study^22^ and an incomplete trial without any results. Another review did not identify any human studies to support the prophylactic use of hydroxychloroquine and chloroquine in COVID-19.^40^ Chowdhury *et* al^38^ was limited to 6 completed studies,^15-17 19 22 28^ and another study with incomplete data.^7^ Another review^42^ was also limited to 7 studies and combined heterogeneous trials with observational studies in a single meta-analysis.^16 22 28 17 27 29 44^ A recent recommendation from the American College of Physicians also concluded that there was insufficient evidence to support the use of hydroxychloroquine or chloroquine for treatment of patients with COVID-19.^43^ Our analysis added additional studies reporting on mortality,^18 21 26^ respiratory support^25^ and QTc prolongation. ^20, 29-34^

During the preparation of the manuscript, we received electronic notification of the publication of two other studies.^44 45^ A observational study of 1438 hospitalized COVID-19 patients from 24 hospitals in New York City (59.7% male, median age 63 years) reported no significant differences in mortality for patients receiving hydroxychloroquine + azithromycin (HR, 1.35 [95% CI, 0.76-2.40]), or hydroxychloroquine alone (HR, 1.08 [95% CI, 0.63-1.85]) compared to controls.^45^ The combination with azithromycin was associated with higher risk of QT prolongation and cardiac arrest. An uncontrolled observational study from the same French hospital with two prior reports,^22 28^ focused on outcomes for 1061 SARS-CoV-2 positive patients treated with hydroxychloroquine and azithromycin and reported that a virologic cure and good clinical outcomes were obtained in 91.7% of patients, with a case fatality rate of 0.96%.^44^ The lack of a control group and the large amounts of missing data preclude any definitive conclusions. A complete evaluation of these studies will be conducted with the next updated search. However, these findings are consistent with our review. Also, this treatment is currently under investigation in several clinical trials and observational studies, which will further clarify the nature of benefit and risk.

Our systematic review has limitations, many of which reflect the quality of underlying data. The methodologic quality of many studies was poor, as noted by others.^46^ Except for a few studies that reported an increased mortality,^18 21^ or no difference in the risk of intubation/death,^26^ the remaining studies were underpowered to detect significant differences in mortality. Few studies reported a formal hypothesis to determine an adequate sample size sufficient to detect the treatment benefit under study and thus may have not detected smaller treatment benefits. A meta-analysis was not feasible because of heterogeneous and novel clinical scales and surrogate endpoints that were not uniform among studies. The interpretation of many of the QT studies was limited by the absence of a control group, assessment of the underlying severity of the illness, and concomitant use of QT prolonging therapies.^18 32^ In addition, the QT interval studies were too small or uncontrolled to establish a risk of life-threatening cardiac arrythmia.

## CONCLUSION

Our systematic review of clinical studies did not provide substantial evidence to support the treatment benefits of hydroxychloroquine or chloroquine in hospitalized COVID-19 patients. Accumulating evidence now raises questions of potential harm including the risk of critical QTc prolongation and increased mortality which deserve further exploration.

## Data Availability

Data will be available on request and are attached as appendix

## ACKNOWLEDGMENT

None

## Supplementary File

Table S1. Search strategy

Figure S1 Quality Assessment of Clinical Trials of Chloroquine/Hydroxychloroquine in COVID-19

Figure S2. Quality Assessment of Controlled Observational Studies of Chloroquine/Hydroxychloroquine in COVID-19

## Contribution

SS drafted the study protocol. Both the authors (SS and TJM) contributed to the search, data extraction, review and interpretation and preparation of the manuscript. SS will act as guarantor of the manuscript.

